# Priorities for Future Research about Screen Use and Adolescent Mental Health: A Participatory Prioritization Study

**DOI:** 10.1101/2021.06.09.21256585

**Authors:** Norha Vera San Juan, Sian Oram, Vanessa Pinfold, Rachel Temple, Una Foye, Alan Simpson, Sonia Johnson, Selina Hardt, Kadra Abdinasir, Julian Edbrooke-Childs

**Affiliations:** Health Service and Population Research Department. King’s College London. UK; McPin Foundation, London. UK; Division of Psychiatry. University College London. UK; Centre for Mental Health, London. UK; Anna Freud National Centre for Children and Families, London. UK; NIHR Mental Health Policy Research Unit. UK

**Keywords:** Screen use, screen time, children, young people, parents, carers, teachers, adolescents, mental health, priority setting, future research

## Abstract

This study aimed to identify research priorities for future research on screen use and adolescent mental health, from the perspectives of young people, parents/carers, and teachers.

**Methods:** The study design was informed by the James Lind Alliance Priority Setting Partnership approach. A three-stage consensus-based process of consultation to identify research priorities using qualitative and quantitative methods. Research was guided by a steering group comprising researchers, third sector partners, clinicians, parents/carers and young people. A Young People’s Advisory Group contributed at each stage.

**Results:** Initial steps generated 26 research questions of importance to children and young people; these were ranked by 357 participants (229 children and young people and 128 adults). Consensus was reached for the prioritization of four topics for future research: (i) the impact of exposure to adult content on young people’s mental health and relationships; (ii) the relationship between screen use and the wellbeing of young people from vulnerable groups; (iii) the impact of screen use on brain development; and (iv) the relationship between screen use and sleep.

Additionally, young participants prioritized questions about online bullying, advertisements targeting young people, and the relationship between social media and specific mental health conditions. Research topics of interest arising specifically during the pandemic included the effects on adolescent mental health of exposure to constant news updates and online racial bias, and how young people take part in activism online

**Conclusion:** These findings will enable researchers and funders to conduct research that is needs-oriented and relevant to the target audience.

Key points and relevance of the study

▪ Evidence about the effects of screen use on adolescent mental health is weak and has been driven by researchers and technology developers.
▪ Young people, parents and teachers prioritized research questions about exposure to adult content online; wellbeing of vulnerable populations; impact of screen use on development; and relation of screen use with sleep.
▪ Young people additionally prioritized research questions related to social media and developing specific mental disorders, online bullying, and companies exploiting adolescents’ vulnerabilities (for example through targeted publicity).
▪ Findings should inform calls for research and funding allocation in order to develop evidence-based policy and guidelines about screen use.

## Introduction

Digital screen use has reached unprecedented levels. This presents opportunities and benefits, such as increased connectivity and access to online mental health support, but concerns have been raised that high levels of usage may harm adolescents’ mental health (Odgers & Jensen, 2020). Recent estimates suggest that 83% of adolescents aged between 12-15 years in the United Kingdom (UK) own a smartphone (Ofcom, 2020). There are widespread reports of increased screen use during the COVID-19 pandemic (Vanderloo et al., 2020). One study of adults (aged 16 years and older) found a 36% increase in screen use during the early stages of the pandemic and lockdown (Rolland et al., 2020). A recent systematic review and meta-analysis of international studies found that for 23% of children and young people smartphone usage was possibly problematic, which was associated with stress, depression, and anxiety (Sohn et al., 2019). Similarly, a systematic review found that social media use may be associated with depression, anxiety, and psychological distress (Keles et al., 2020).

However, existing evidence does not provide clear support for the hypothesis that screen use causes mental health problems. A recent review found that the length of time young people spent using digital media was not consistently associated with increased mental health difficulties (Odgers & Jensen, 2020). Another recent systematic review found moderate evidence of a negative association between screen time and some health outcomes, although the number of studies specifically examining smartphones was limited. This raised questions about the relevance of recent evidence to young people since they are predominantly smartphone users (Stiglic & Viner, 2019). These studies have largely investigated about associations between quantity of screen time and mental health outcomes. This has been criticised as a reductionist approach that does not consider type and context of screen use. There have also been calls for research to consider the potential benefits of screen use (Prinstein et al., 2020).

Increasing the relevance of future research requires that young people be engaged in the identification of research priorities. This is more important now than ever given increased reliance on screens for education, social interaction and service access due to the COVID-19 pandemic, especially given recent evidence suggesting rises to 1 in 6 young people aged between 5 and 16 experiencing mental health disorders (NHS Digital, 2020).

The extent to which screen use has a role in mental health—and what this role is—is uncertain, limiting the development of evidence-based policy and practice, despite significant policy interest in the UK (*Internet Safety Strategy Green Paper*, 2018; *Online Harms White Paper - April 2019 - CP 57*, 2019; *Transforming Children and Young People’s Mental Health Provision: A Green Paper*, 2017) and elsewhere. A crucial first step to the development of meaningful evidence-based policy and guidelines and the effective use of resources is ensuring the right research questions are being asked (Terry et al. 2018). Yet to date, the research agenda in this field has been mainly driven by the research community, technology developers, and policy makers, with little input from young people, parents and carers, or teachers (Hollis et al., 2018). The aim of this study is to address this gap.

### Aims

We aimed to identify the top research priorities regarding screen use and young people’s mental health from the perspective of young people aged 11-25 years old, parents/carers, and teachers.

## Methods

Our study design was informed by the James Lind Alliance Priority Setting Partnership approach (James Lind Alliance Priority Setting Partnerships, 2018). Accordingly, research priorities for screen use and young people’s mental health were identified through a three-stage process of consultation and consensus (see Figure 1).

**Figure 1.**
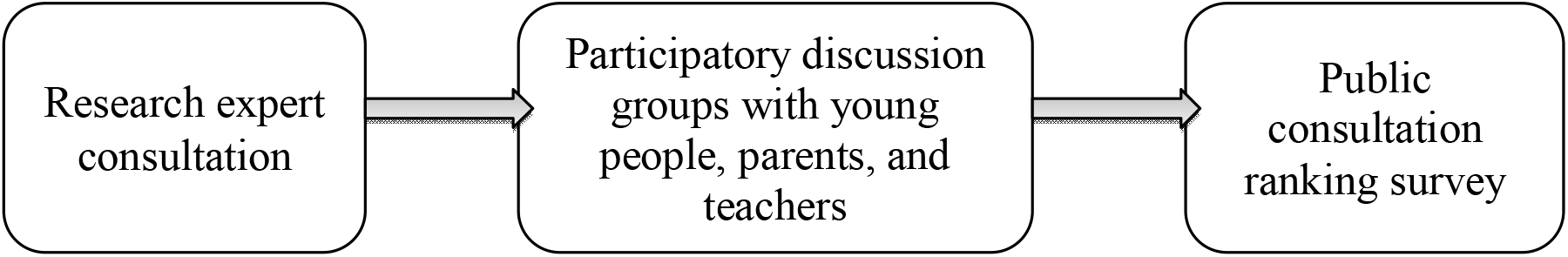
Flow chart presenting the three-stage process of consultation and consensus

This process was guided by a Steering Group formed by researchers, clinicians, voluntary sector partners and parents/carers, and young people. They met bimonthly to discuss recruitment, data collection, and interpretation of findings. The Young People’s Advisory Group (YPAG), part of the Young People’s Network coordinated by McPin Foundation, were a central part of the process, meeting with the research team regularly and feeding into all work stages including study design, promotional work and write up (Sellars et al., 2020).

In this study, we focus on recreational and personal screen use. We define this as watching TV programmes and videos, playing games, using social media, or browsing the Internet excluding time spent for education, employment, or training purposes. We define social media as “Internet-based channels that allow users to opportunistically interact and selectively self-present, either in real-time or asynchronously, with both broad and narrow audiences” (Carr & Hayes, 2015, p.50)

The University College London research ethics committee approved this study (Project ID: 14037/001).

### Step 1 Preliminary consultation to develop a long list of relevant research questions

We drew on a previous research priority-setting study focused on young people’s mental health in general (McPin Foundation, 2018). We extracted from these 66 questions relevant to screen use and we contacted researchers with relevant expertise and asked them to: (1) identify research questions omitted from this list but which they perceived to be important, and (2) advise us of research that was underway to address the existing questions. Respondents could provide this information either via an online questionnaire or in a telephone call with the research team.

### Step 2 Participatory group discussions to build on and refine the long list

The next step involved participatory discussion groups with stakeholder groups to build on and refine the long list of questions from Step 1. A more detailed description of the methods for this step can be found in a companion paper (Vera San Juan et al., In preparation).

### Recruitment and data collection

Participatory discussion groups were advertised through the Steering Group’s networks. Groups were held separately for young people (aged 11-25 years old), parents, and teachers. Due to the introduction of lockdown measures in the UK as part of the response to COVID-19 in March 2020, three of the 12 planned groups could not take place and participants instead received an online survey covering the same discussion topics.

Discussion groups included three main parts: (1) individual reflection, where participants were asked to reflect and write about their screen activities and how these made them feel; (2) pair work, where participants discussed their answers to part one, developed a mind map to describe their experience of using screens and formulated three research questions that they considered important; and (3) group discussion, where pairs presented their mind map and research questions to the group and agreements and differences were discussed. Facilitators also explored the groups’ views about the importance of questions from the existing list from McPin Foundation and expert consultation. The topic guide is provided in Supplementary Material 1. The YPAG contributed significantly to the development of all recruitment and data collection materials. Additionally, group discussions with young people were co-facilitated by NVSJ and a young peer researcher, supported by Centre for Mental Health, to ensure adequacy for the younger participants.

Discussion groups were audio recorded and suggestions for specific research questions from each pair collated. Participants were also given the option to leave research questions anonymously in a box at the end of the session.

We anticipated that rich data would be collected from eight focus groups with young people with and without self-identified experience of mental health problems; four groups of parents/carers; and two groups with teachers. Emphasis was placed on geographical spread in England holding groups in a range of locations, with a mix of gender and ethnicity.

### Analysis

Analysis was conducted applying the Rigorous and Accelerated Data Reduction (RADaR) technique (Watkins, 2017). This is a rapid matrix-based content analysis method used to collaboratively reduce raw qualitative data into a final set of project deliverables (Vindrola-Padros, 2020). NVSJ and JEC made notes and transcribed relevant quotes from the discussion group recordings onto a Microsoft Excel matrix where rows were cases (each group) and column headings included: *Proposed research questions; key positive and negative aspects of Screen use; key positive and negative aspect of social media; key positive and negative aspects of gaming*. NVSJ and JEC discussed the themes emerging in each row and compiled them in a final *Main topics* column. These emerging themes were reviewed again in a final data reduction cycle with UF and the YPAG, where themes were formulated as research questions.

### Step 3 Public consultation for final prioritization

#### Recruitment and data collection

A long list of 200 questions was generated combining questions identified in (1) the McPin Foundation JLA priority setting partnership (McPin Foundation, 2018); (2) our step 1 expert consultation; (3) our Step 2 discussion groups; and (4) Steering Group and YPAG meetings. Through two more RADaR data reduction cycles (Watkins, 2017), the Steering Group and YPAG grouped similar questions and excluded questions deemed out of scope. Care was taken to ensure the wording of the questions going forward to the next phase reflected that of the original questions. The resulting list of 26 research questions was included in an online ranking exercise conducted with young people, parents/carers, and teachers.

After providing informed consent, participants were asked to drag and drop the ten questions most important to them from the list of 26 and then rank these ten questions in order of importance (see figure 2). The 26 questions were presented in an aleatory order for each participant. The survey also included questions about demographic characteristics, such as gender, ethnicity, region, and experience of mental health difficulties, and an additional open-ended question about new research topics of interest due to COVID-19 isolation measures.

**Figure 2.**
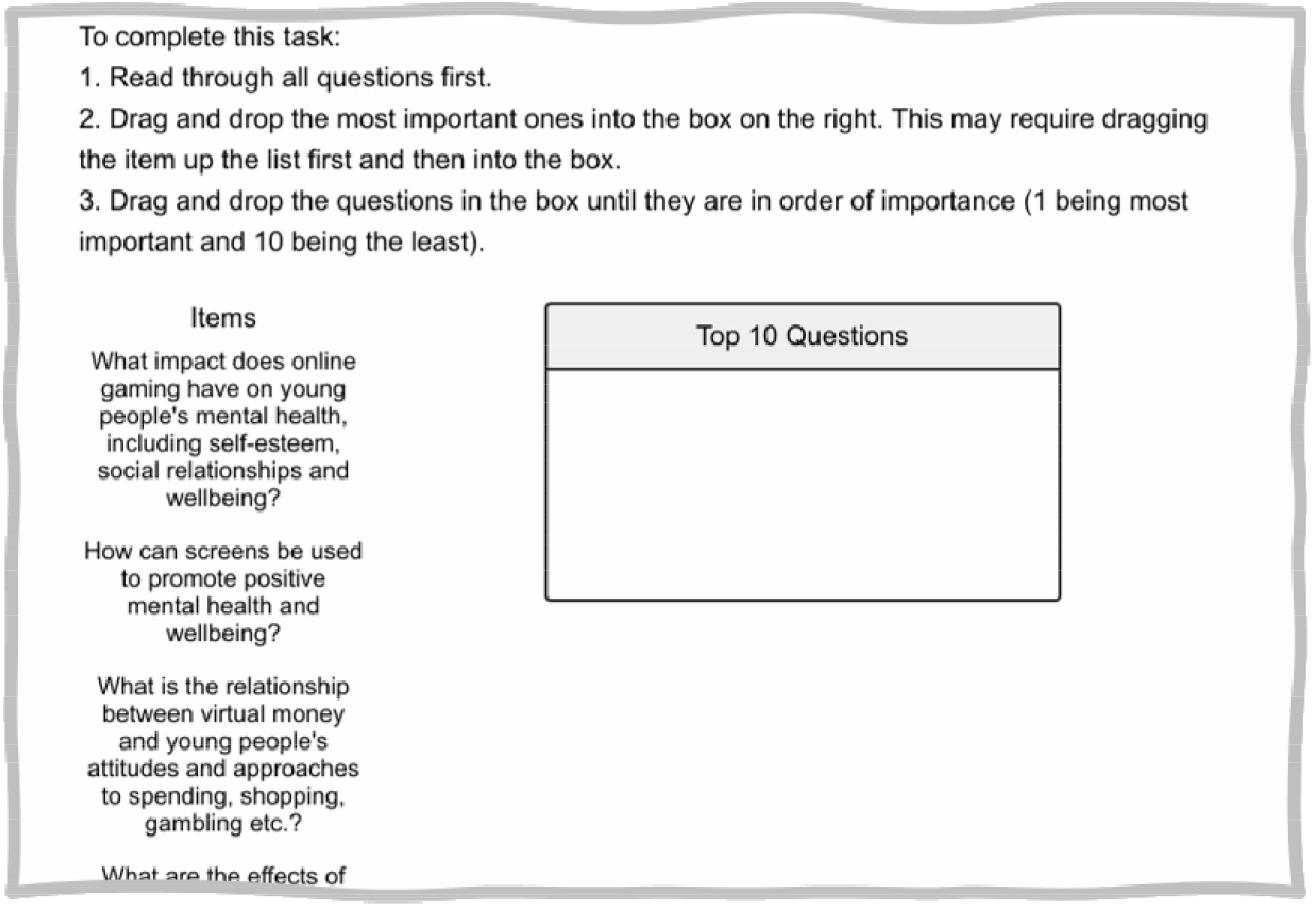
Ranking survey task.

Aiming to achieve rapid recruitment of a large and varied sample, the survey link was disseminated through the Steering Group networks via email, Twitter, Facebook, Instagram and blogs on pertinent websites. Additionally, we used a recruitment company (<u>Panelbase</u>) to increase recruitment of young people during lockdown. Recruitment materials specifically targeting younger demographics were co-developed with the YPAG and young peer researchers and used on social media.

### Analysis

Survey results were extracted into Microsoft Excel. Results were analyzed for the full sample and by subgroup: young people under 16 years old, young people 16-25 years old, young people with lived experience of mental health problems, young people without lived experience, and adults (parents/cares and teachers).

The main indicator of importance was the number of times a question had ranked within the top ten priorities list. Other indicators were explored and considered in Steering Group discussions, such as the number of times a question was ranked as 1^st^ most important or the average position in the ranking.

The open-ended question about changed perceptions due to COVID-19 lockdown measures was explored first using word frequencies to identify the main topics arising, followed by NVSJ’s review of all answers to identify research questions proposed in relation to these topics.

The final top research priorities were reviewed for clarity by the YPAG and Steering Group, prior to study completion.

## Results

### Sample

#### Step 1 Preliminary consultation to develop a long list of relevant research questions

Nine experts added relevant questions to our lists on topics including: impact of poor internet access and social media on vulnerable populations; the effects of constant surveillance on adolescents’ development; and strategies to bring together adults and young people’s perspectives on screen use.

#### Step 2 Participatory discussion groups to build on and refine the long list

We conducted a total of 12 discussion groups with 4-8 people (total N=68; seven groups of young people, N=46; three groups of parents/carers, N=15; and two groups of teachers, N=7). Each lasted approximately one hour and took place between January-March 2020 in Greater London; East Midlands; Yorkshire; and South West England.

Qualitative findings are presented below alongside the list of top research priorities identified in Step 3.

A detailed description of the sample is presented in Table 1 and the above-mentioned companion paper (Vera San Juan, In preparation).

**Table 1.** Discussion group sample description

|  |  | **N (%)** |
| --- | --- | --- |
| **Ethnicity** | White British/European | 44 (65%) |
|  | Other | 20 (30%) |
| **Gender (N, %)** | Female | 45 (66%) |
|  | Male | 22 (33%) |
| **Mean age (SD, range)** |  | 20 (11-56) |
| **Participants with experience of mental health problems (N, %)** | Current | 20 (30%) |
|  | Past | 20 (30%) |

*Note.* Total N = 68.

#### Step 3 Public consultation for final prioritization

Data were collected from 22^nd^ June to 20^th^ September 2020. A total of 822 people clicked on the survey link, of whom 357 completed the main ranking task (229 young people; 128 adults) and 330 (210 young people; 120 adults) answered the open-ended questions. Attrition was greatest among parent/carer and teacher participants (48% of those who started the survey did not complete it).

A detailed description of the sample is presented in Table 2. The sample was predominantly white (N=279; 78%) and resident in England (N=312; 87%). Half lived in cities/suburban areas (N=179; 50%). The mean age of young people was 19 (range 11-25) and of adults 43 (range 21-63). Among young people, 55% (N= 127) reported having experienced mental health problems currently or in the past (most commonly depression, anxiety, or eating disorders) while among adults 54% (N= 70) reported that they or their child experienced mental health problems currently or in the past (most commonly anxiety, depression, and self-harm).

**Table 2.** Public consultation survey sample description

|  |  | **N (%)** |  |
| --- | --- | --- | --- |
| **Region** | England | 312 (87%) |  |
|  | Scotland | 22 (6%) |  |
|  | Wales | 18 (5%) |  |
| **Ethnicity** | White British/European | 279 (78%) |  |
|  | Asian/Asian British | 37 (10%) |  |
|  | Black/African/Caribbean/Black British | 16 (5%) |  |
|  | Mixed/Multiple ethnic group | 20 (6%) |  |
|  |  | **Young people** | **Adults** |
| **Gender (N, %)** | Female | 140 (61%) | 91 (71%) |
|  | Male | 86 (28%) | 36 (28%) |
| **Mean age (SD, range)** |  | 19 (11-25) | 43 (21-63) |
| **Participants with experience of mental health problems (N, %)** | Current | 86 (38%) | 40 (31%) |
|  | Past | 41 (18%) | 30 (23%) |

*Note.* Total N = 357.

### Priorities for research about screen use and adolescent mental health

Across the various subgroups, participants agreed on four priority research questions. These are presented in Box 1 and explored below alongside related qualitative data to contextualize what the question meant and why it was considered important, collected across stages 1, 2 and 3. Differences between groups and findings specifically emerging in groups of people with lived experience of mental health problems are highlighted. A full list of the 20 top research priorities identified across all participant groups can be found in the Supplementary Material 2, disaggregated by young people, those with and without mental health problems, and adults.

#### Box 1.

Research questions ranked as top research priorities for research across subgroups (young people with and without lived experience of mental health problems, parents/carers and teachers).

▪ What impact does exposure to adult content (e.g., violent, sexual) have on young people’s mental health and relationships to others?
▪ What is the relationship between screen use and mental health and wellbeing for young people from vulnerable groups (e.g., mental or physical health conditions, disability, learning difficulties)?
▪ What is the impact of screen use on brain development?
▪ What is the relationship between screen use, sleep and mental wellbeing in young people?

#### What impact does exposure to adult content (e.g., violent, sexual) have on young people’s mental health and relationships to others?

Participants across groups were interested in impacts of exposure to “adult” content. Young people specified certain types of content as especially potentially harmful; they tended to see violent video games as largely harmless and oriented to developing skills and a sense of achievement, watching “gore videos” intended to test tolerance of violence to the limit caused greater concern. Some also reported encountering unwelcome content when following automatically suggested content online. Most apps offered ways to report and block content that seemed inappropriate. However, young people were concerned that content censoring relying on individuals could minimize the perspectives of people with lived experiences. For instance, people without knowledge of mental health conditions may report photos of healed self-harm scars as violent content, even though these photos were meant to encourage others to recover.

> Young person, Stage 1 focus group. London *“fully healed scars are often flagged because they’re self-harm, but people might not mean it as gore-y, they want to show they’ve recovered. then people post things about alcohol and taking drugs and that isn’t classified as self-harm, they’re just having fun”*

Teachers highlighted that the ease with which young people shared images and increased accessibility of pornographic material had increased sexualization of thinking and behavior of young people. Parents/carers were frustrated by the lack of control they had over children’s exposure to online content. This included pornographic material and distressing constant news updates about phenomena such as global warming or COVID-19 news during lockdown, which blurred the lines between normal educational content and potentially distressing content. Conversely, positive initiatives and good news had been important motivators and encouraged participants through difficult periods during the lockdown.

> Parent/carer, public consultation. Midlands *“In some ways it [internet] has helped my daughter to keep connected with friends to overcome some of her anxieties, helped her to keep her mind occupied. […] On the other hand, she has been spending too much time in front of a screen, exposed to more news about the virus at times without her wanting to which has increased her anxiety*.*”*

Young people underscored the importance of making online a safe and positive space to grow during the pandemic as it was *“all us teens have right now”* (Young person, public consultation. Midlands).

#### What is the relationship between screen use and mental health and wellbeing for young people from vulnerable groups (e.g., mental or physical health conditions, disability, learning difficulties)?

Young people mentioned that online communities made them feel understood, however, people feeling isolated or vulnerable might inadvertently engage with online communities that had the potential to cause harm. Participants commented on their experiences with eating disorder communities saying that they very quickly saw themselves immersed in uncomfortable conversations with “Ana coaches” (anorexia coaches) and did not know how to undo sharing images or information they had revealed before realizing it was inappropriate. Accordingly, young people with lived experience ranked highly the question: *“What are the pathways that lead adolescents to websites and blogs that promote harmful behaviors and what is the impact on their mental health?”*.

> Young person, group discussion. Yorkshire *“[about anorexia communities] It’s people understand how you feel, whereas no one else understands that. And, like, you have one goal and other people are trying to help you on this goal. So, you think it may be bad… but they’re helping you”*

The anonymity of bullies online in these or other online contexts was thought to be a key barrier to solving the problem. Young people from vulnerable groups felt there was nothing they could do against bullies online because their real identity would never be revealed and therefore, they would not be punished.

Researchers participating in our stage 1 consultation raised concerns about young people with vulnerabilities spending more time on screens due to their accessibility in comparison with participating in activities outdoors. Researchers and young participants in discussion groups also voiced their concerns about publicity targeting vulnerable populations and referred to algorithms behind online advertising as “*covert social sorting*”. Examples of this were gambling adverts targeting youth from low-income families, and adverts for diet products targeting young women. Young people between 16-25 years of age ranked this topic in their top-ten priorities for future research.

> Young person, discussion group. London *“In real life young people wouldn’t go to a casino or a betting house… [but], there are a lot of adverts online and it only takes one click to get in”*

Another concern of economically disadvantaged young people with poor quality broadband, devices, and content. Adult participants mentioned that the reliance on screens during lockdown would perpetuate educational disparities and thus hinder future opportunities for young people.

> Parent, London *“it’s highlighted the educational disadvantage children are at if the only device they have available is a phone (it’s very hard to complete GCSE work on one)”*

Conversely, participants mentioned more stimulating online content had become available during lockdown due to artists, theatres, galleries and workshop platforms sharing content for free.

#### What is the impact of screen use on brain development?

This question was interpreted in relation to the development of cognitive and social skills, not necessarily structural development of the brain. Numerous screen activities which facilitated the development of new skills were mentioned by young and adult participants, such as learning and sharing on YouTube and online forums.

> Parent, London *“[talking about YouTube] Young people are empowered because they can learn new skills and take on challenges that build confidence”*

Young participants wanted to know about the effects of socializing mainly online vs. having interactions in person. Adults thought interaction through screens was hindering young people’s development of empathy, communication skills and attention, though opinions became more favorable during lockdown.

Parents expressed concern over children constantly multitasking and not living in the moment. Examples of this were constant texting while doing activities or recording everything to post it later.

> Parent, discussion group. Yorkshire *“Because they’re talking at each other, not with each other. […] They probably haven’t said, “How are you today?” or “What have you done yesterday?” […] You look at their conversations, but they’re not conversations even through WhatsApp or messages or whatever. And they’re not – they don’t make any sense*.*”*

Teachers also reported an increase in attention difficulties and attributed this to children being used to content that was excessively stimulating and quick. Teachers suggested that young people were not developing patience and other necessary skills to cope with failure due to gaming providing instant gratification (easy wins) or the possibility to start over until they won.

> Teacher, discussion group. Yorkshire *“Now the pace of the classroom isn’t fast enough for them. It’s like they constantly want to be done, done, done, done, done, done, done. And you ask them to do any lengthy task where they have to actually sit back and stop for a minute. They have a meltdown”*.

Some young people commented that parents’ excessive use of screens affected young people’s development as parents interacted less with their children. Parents being “*hooked*” on apps distracted their attention from their children’s wellbeing. Complementing this, the effects of parental screen time on young people’s mental health was ranked by adult survey participants as one of their top-ten priorities for future research.

> Young person, discussion group. East Midlands *“when parents spend way too much time on their screens, like, whether it’s working, or on social media, and they neglect their child due to social media. Because I know my mum used to do it, where she’d be on Tinder and stuff like that, that much she’d forget I was even there”*

#### What is the relationship between screen use, sleep and mental wellbeing in young people?

Young people participating in this study reported apps with *“endless scrolls”* as the most time-consuming and ultimately tiring, and suggested that using these apps first thing in the morning and at bedtime could be problematic. This concern was reflected in the question “*Do companies exploit addictive behaviors (e*.*g*., *games, online gambling, algorithms behind social media and targeted publicity)?*” which young people ranked among their top 10. Teachers, however, perceived that gaming caused the most disturbances to sleep patterns and mentioned having seen children exhausted during the school day due to playing games through the night.

Young participants, particularly those with personal experience of mental health problems, highlighted positive night-time screen routines such as spending time on meditation apps and ending the day with positive texts exchanges with their support groups. They described as problematic the practice of parents confiscating their phones at night without taking into consideration possible positive uses.

> Young person, discussion group. East Midlands. *“there should be a negotiable control over phones. Because I got into some pretty deep water with my mental health a couple of months ago, and my parents, […] freaked out and put away all my electronics… and that made me feel worse. They were convinced that I was talking to people at night. And I’m, like -Night time is when is I need my phone so that, if I’m up because I’m having nightmares…”*

### Research interests during lockdown

Most of the participants believed that the questions included in the public consultation (Step 3) covered their main research interests, suggesting that the pandemic had not substantially changed views on research priorities. Perceptions about screens generally became more positive, with adults becoming more interested in understanding young people’s online activities and acknowledging their expertise. They particularly reflected on the important role of screens on young people’s education and the potential of screens as a tool to empower.

> Teacher, public consultation. London *“What strategies (if any) are young people using to manage their own screen use and balance with non-screen-based activities?”*

A new research topic that gained prominence was how young people used the Internet to learn about different cultures and participate in activism. Participants proposed looking into online racial biases and social change through online movements as a new research topic, introducing new questions such as *“How screen time has enabled young people to educate themselves and partake in activism during COVID19?” (Parent, public consultation. London)*.

> Parent, public consultation. London *“They [screens] have been the only social interaction my children have been able to have. They have educated themselves about BLM [Black Lives Matter] and LGBTQ+ issues through social media, their social networks have broadened, and they are friends with other young people around the world*.*”*

## Discussion

### Principal Results

This study identified future research priorities on young people’s screen use and mental health based on a three-step process, from the perspective of young people, parents and carers, and teachers. From a list of 26 important research questions that were included in the final public consultation, adults and young people agreed on four. The topics covered in these questions were: exposure to adult content online; wellbeing of vulnerable populations; impact of screen use on development; and relation of screen use with sleep.

Young participants agreed on an additional three research questions related to social media and developing specific mental disorders, online bullying, and companies exploiting adolescents’ vulnerabilities (for example through targeted publicity). While parents and teachers expressed specific interest in the effects of screen use on adolescents’ ability to maintain attention, the effects of parental screen use on young people’s development, and ways to support young people for adequate screen use.

Participants’ views on screen use and young people’s mental health became more favorable during lockdown, potentially due to greater reliance on screens for education, communication, and entertainment and other significant events including the Black Lives Matter movement. Research topics of interest arising during this time included the effects on adolescent mental health of exposure to constant news updates and online racial bias, and how young people take part in activism online.

### Comparison with Prior Work

Research priorities identified in this study corresponded with previously identified evidence gaps (Hollis et al., 2018; Odgers & Jensen, 2020; Ofcom, 2020), including: (1) the relation between social media and specific mental disorders; (2) disentangling the use and impacts of different types of screens (individual vs. social, or work vs. leisure, etc.); (3) learning from adolescents about positive uses of screens; (4) tending to the digital needs of vulnerable populations; and (5) comparisons between online and face to face interactions.

The YPAG advising this study highlighted the need to focus on screen “use”, rather than screen “time”, and the blurring between the use of screens for educational and leisure purposes. The limitations of basing observations based on “time” rather than the activity or context in which screens are used has also been discussed elsewhere (Ellis, 2019; Odgers & Jensen, 2020; Prinstein et al., 2020). A review by Verduyn et al. (2017) found “passive” and “active” usage of social network sites had different consequences for subjective wellbeing. Passive use provokes social comparisons and envy, while active use created social capital and stimulated feelings of social connectedness.

Recent reviews have pointed to a lack of evidence for an association between the amount of time that adolescents spend on screens and poor mental health (Kaess, 2020). Research to date is mainly cross-sectional, and therefore correlational, and cannot be used to infer whether screen use leads to mental health problems or whether young people with existing vulnerabilities are more likely to use screens in unhealthy ways (Odgers & Jensen, 2020). Two recent longitudinal studies found that only excessive amounts of social media use were linked to mental health problems (Riehm et al., 2019; R. M. Viner et al., 2019), and this was likely mediated by experiences of cyberbullying, little physical activity, or poor sleep (R. M. Viner et al., 2019). Another longitudinal analysis of nationally representative samples in Ireland, United States, and United Kingdom found a small significant negative association between technology use and well-being (Orben & Przybylski, 2019). However, Foster & Jackson (2019) argue that the very large number of people engaging with technology warrants considering even small negative links with mental health.

In this study, participants expressed concern that social skills development would be hindered by interacting with others online rather than in person. Evidence on this point is unclear. For example, Downey & Gibbs, (2020) compared teachers’ and parents’ evaluations of children’s social skills in two cohorts between 1998 and 2010. their ratings suggested social skills had not changed in the more recent cohort, even when accounting for sociodemographic factors and screen time use. Our study findings highlighted adults’ increased awareness of the positive social aspects of screens during lockdown; similar findings were reported by Ofcom (the UK’s Office for Communications) in a recent COVID-19 review (Ofcom, 2020).

There is thus a need to better understand positive uses of and impacts of screen use. Simultaneously, more work is needed to understand individual vulnerability or resiliency factors that may impact online experiences: Research suggests that online risks are likely to mirror offline risks, and has drawn attention to the lack of support for young people struggling in either sphere (Hollis et al., 2018; Kowalski et al., 2014). Additionally, as suggested by the Royal College of Pediatrics and Child Health, parents and young people are experts by experience and should have active input in screen use guideline development (R. Viner et al., 2019).

#### Strengths and limitations

This project combined strengths of qualitative and quantitative research. The mixed methods approach allowed both the identification of research priorities and an understanding of people’s perspectives on the prioritized topics. Results were also enhanced by multiple stakeholders, including – crucially – young people collaborating across all stages of the work.

The COVID-19 pandemic began during our study. Although, when asked in the public consultation survey, participants did not appear at the time to indicate that their views had substantively changed, it is important to recognize this major change in global context. The long-term impact of different patterns in screen use due to the pandemic and restrictions may have increased the ranking of certain research questions. Due to the national lockdown, we did not achieve our target sample size and diversity for the step 2 discussion groups. However, the existing sample was considered to have sufficient information power to achieve the research objectives. The research questions targeted in discussion groups were specific and groups focused solely on addressing them (Malterud et al., 2016).

To the best of our ability, and with input from our YPAG, we created a safe environment for participants to discuss their priorities within a group setting. There are, however, topics l that people are likely to find uncomfortable to talk about, for example access to pornographic material, which did not come up in the expert consultation or group discussions.

Finally, ranking surveys are subject to potential lack of reliability. Empirical results have suggested the stability of ranking information decreases with decreasing rank (Ben-Akiva et al., 1992). To mitigate this potential limitation, we designed a two-step task for people to select their top priorities and combined the indicators that were thought to be most reliable. Almost half of adults who started the survey did not complete it. Feedback suggested this was due to increased workload during lockdown and poorer survey functionality on mobile phones. The limited participation of parents with higher workloads or poorer access to devices may have affected the range of parent’s research interests identified in the study.

#### Conclusions

Our collaboration with young people and focus on young peoples’ views allowed us to identify research priorities on screen use and adolescent mental health, and to gain insight into the reasoning behind these research questions. Recent events have sparked special interest in the effect of screen use on young people’s mental health. This work points to the great need for more evidence in this field and potential risks and benefits at stake. Findings should inform calls for research and funding allocation in this field in order to develop evidence-based policy and guidelines.

**Lived experience commentary written by Syinat Tageava. Member of the YPAG**

As a young person, it is great to see that young people were involved at every stage of this research, however, it has some drawbacks. There are clear points in the paper that would have been hard to see without direct input from young people. For instance, the high number of dieting advertisements that young girls see. I was glad to see the significance of online activism highlighted by this paper, as news of breaches of human rights continues to be shared daily. I was also pleased to see that parental screen time came up as a topic for future research, since it is a much overlooked factor in young people’s mental health.

I hope that future research, perhaps based on the sub questions provided by this paper, makes it a priority to have more male responders. This paper seems very one sided since only 28% of young participants identified as male. It is disappointing to see the mostly negative stereotypical impacts of exposure to adult content. Positive impacts such as sexual education and empowerment were not mentioned, possibly because responders were uncomfortable sharing this information. I have found that the internet is an open platform for various people to honestly communicate, so young people can find empowering support groups (e.g. sexual assault survivors) and explicit sexual health advice they cannot find in real life. Young people can also learn about issues such as understanding how to give consent; and the legal definitions involved in situations of a sexual nature.

Overall, this paper has brought to light some crucial questions, that can inform future research. Preferably, this research will have just as much involvement from young people.

## Supporting information

YPAG Consent

YPAG Consent

## Data Availability

Data relevant to the study are included in the article or uploaded as supplementary information. Further information can be requested by contacting the corresponding author.

## Abbreviations

YPAG: Young People Advisory Group
RADaR: Rigorous and Accelerated Data Reduction technique

