## Supplementary material 1 and 2 for "Priorities for Future Research about Screen Use and Adolescent Mental Health: A Participatory Prioritization Study"

### Supplementary material 1 – Discussion group topic guide

**Topic Guide for Small Group Discussions/ Focus Groups**

***We want to discuss with young people, parents/carers and teachers their views of the benefits of recreational screen use and any concerns they share, linked to mental health.***

Key considerations:

- The topic guide will be tailored to the needs (e.g., age) of the group.
- It will be lead by participants and their views.
- Facilitators will keep an eye out for anyone who may be feeling uncomfortable and will refer them to the support mentioned in the information sheet if needed.

***Use of this guide***

This topic guide is planned as a starting point for a discussion with young people (11-25 y/o), parents/carers and teachers about the important questions and key areas of interest regarding young people’s mental health and recreational screen use. The topic guide should provide an overall structure of the discussion to be covered. The probes and general direction of the group will also be informed by participants’ reflections and concerns, and questions will follow participants’ interests and any points that need further clarification.

***Materials needed for each session***

- Name labels (sticky).
- Small soft ball for introductions.
- Pictures of different types of “screens” (phone, laptop, tablet computer), emojis, and different social networks or apps such as Snapchat, Instagram, TV, WhatsApp, YouTube, etc. to help participants develop mind maps during group activities.
- A4 papers and pens for each participant to write down their thoughts regarding screen use and mental health (individual work).
- Flip chart paper, pens and post-it notes for participants to work in pairs to describe benefits and/or any concerns/disadvantages of screen use that they share.
- A6 landscape cards to write each topic of interest on one side and reflections or notes for clarification on the reverse.
- Board and yarn to pin/stick topic cards and create a final theme map.
- Empty box for anonymous comments/suggestions

***Set up considerations***

We will work with local leads who help us set up the groups to ensure we have considered the following:

- Room layout and adequate lighting/heating.
- Refreshments available.
- Facilitation team know roles and who will support a young person if they want to leave the group or become distressed.
- We will ask the link person at the organisation if any participants have special needs.
- Other health and safety considerations (e.g., toilets, fire escapes).

***Session overview***

- Introduction
- Individual reflection
- Working in pairs
- Group discussion
- Wrapping up

***Part one: Introduction (10 minutes)***

- Facilitator introduction.
- Link person from host organisation introduces facilitator (and possible co-facilitator).
- Short study introduction: Based on information sheet – aims of the study; what’s going to happen during small group discussion; duration; reminder of recording and use of data, confidentiality and right to pause or withdraw. Young people will be reminded that their answers will not be shared with their parents or teachers unless it is an emergency.
- Ground rules for the discussions: All views are welcome and there are no right or wrong answers. Listening to and respecting other people’s ideas, only one person should speak at a time, you may share news of this event with other people but not sharing specific responses or ascribing individual names to specific statements. Ask people if they propose additional rules.
- Group member introductions: each person says their name, what’s the last show they watched and what social media they use most. We may do this as a game (if the group wishes) with a ball and people throw to the next person to introduce.

***Part two: Individual work (10 minutes)***

To be answered on the A4 paper individually. We will ask people to make notes and say the next exercise will be to comment on32

. these notes working in pairs. Questions will also be printed or projected so they are available at any time.

1. What types of screens do you use in your free time? [use pictures of different types of “screens” as a stimulus]?
2. What do you do on each device? [use pictures of social media/films… as a stimulus]. Prompts: Do you use this screen on your own, or with friends/family? Are there things you do much more than others?
3. How does using these screens make you feel? Prompts: What do you get from using these screens? In what way does it have a good/bad effect on you?

Prompt if necessary: We are talking both about mental health and mental health problems (feeling good, like being happy and entertained, and connecting with others, and feeling bad like feeling lonely, ashamed, anxious or stressed), so both positive things and challenges.

***Part three: Pair work (25 minutes)***

- In pairs (or groups of three), share with each other what you have written and discuss how these topics are related. Use the flip chart and other materials available to portray different experiences and ideas around screen use and your feelings and behaviours  (mental health) in a mind map format (examples of what a mind map is will be provided).

With this “map” in mind, consider in pairs:

1. What are the important questions/ topics about  screen use and mental health that you think research could answer? [Alternative wording: What would you like research to find out about in relation to the things you included in your mind map?/ What do you think people should be researching or trying to understand better?]
2. Write on the A6 cards one topic per card (3-5 cards in total). On the back of the card write any reflections or notes about this question/ topic. [In the case of the younger groups, the moderator will help by writing out the pair’s main ideas.]

Questions will be printed or projected. Facilitators will go around the room to address any questions. A brief break will take place here and refreshments will be offered.

***Part four: Group work (30 minutes)***

Group discussion will be encouraged in a way that participants feel comfortable and sharing with the group feels natural and safe.

1. Each pair presents their A6 cards with the topics/questions to the group. Pairs contribute with their views on these topics/questions and the reflections behind their choice referring to their flipchart when useful.
2. After each pair presents their topics/questions, they hand the A6 cards to the facilitator. Facilitator will encourage whole group discussion to begin laying out cards on the board roughly mapping relations and overlap between topics.
3. Once all pairs have presented and their cards are placed on the board, the “mind map” generated will be discussed again by the group and edited as they suggest. Agreements and differences that may have arisen will be considered. The soft ball may be passed around to encourage all to participate.
4. The facilitator will assess whether prevalent topics/questions from other sources have not come up in this group discussion and ask for the group’s opinion about them.

***Part five: Wrapping up (10 minutes)***

Participants will be thanked for their time and hard work.

- Do you have any comments about screen use and mental health that you would like to add?
- Do you have any questions about the research or the use of the information?

The moderators will remain in the room for an extra 10 minutes and remind participants of the contact details and participants will be told they can ask them any additional questions or disclose anything they preferred not to mention in front of the group.

Participants will be asked to fill in a form with their contact details so we can keep you informed of the progress of this study and feedback on the group discussion experience.

### Supplementary Material 2 – List of 20 most important questions

List of important questions about screen use and adolescent mental health

| **Screen use and adolescent mental health**  **20 most important questions** | **YOUNG PEOPLE*** | **WITH LIVED EXPERIENCE** | **WITHOUT LIVED EXPERIENCE** | **ADULTS** |
| --- | --- | --- | --- | --- |
| What impact does exposure to adult content (e.g., violent, sexual) have on young people's mental health and relationships to others? | ✓ | ✓ | ✓ | ✓ |
| What is the relationship between screen use and mental health and wellbeing for young people from vulnerable groups (e.g., mental or physical health conditions, disability, learning difficulties)? | ✓ | ✓ | ✓ | ✓ |
| What is the impact of screen use on brain development? | ✓ | ✓ | ✓ | ✓ |
| What is the relationship between screen use, sleep and mental wellbeing in young people? | ✓ | ✓ | ✓ | ✓ |
| Does social media and screen use increase specific mental health problems in young people, such as self-harm or eating disorders? | ✓ | ✓ | ✓ |  |
| What is the impact of online bullying or trolling on young people's mental health? | ✓ | ✓ | ✓ |  |
| Do companies exploit addictive behaviours (e.g., games, online gambling, algorithms behind social media and targeted publicity)? | ✓ | ✓ | ✓ |  |
| Do young people become addicted to screens and if so, is it similar to other forms of addiction? | ✓ |  | ✓ | ✓ |
| What are the disadvantages of not having access to screens/internet for young people from poorer backgrounds? | ✓ | ✓ |  |  |
| How does online schooling affect adolescent's mental health? | ✓ |  | ✓ |  |
| What impact does online gaming have on young people's mental health, including self-esteem, social relationships and wellbeing? | ✓** |  |  |  |
| How can screens be used to promote positive mental health and wellbeing? |  | ✓ |  |  |
| What are the pathways that lead adolescents to websites and blogs that promote harmful behaviours and what is the impact on their mental health? |  | ✓ |  |  |
| What is the relationship between online platforms (e.g. YouTube) and the development of skills, confidence and empowerment in young people? |  |  | ✓ |  |
| Does screen use make people feel more or less connected with others than real life interactions? |  |  |  | ✓ |
| What is the impact of online communication on young people's development of communication and relationships skills? |  |  |  | ✓ |
| Does screen use interfere with young people's capacity for attention? |  |  |  | ✓ |
| How can adults support young people to manage screen use? |  |  |  | ✓ |
| What is the impact of parental screen use on young people's mental health? |  |  |  | ✓ |
| What is the relationship between increasing surveillance of young people and their mental health? *** | ✓*** |  |  |  |
| * weighted average between <16 and 16+ age groups | | | | |
| ** among top 10 important question for young people < 16-years old only | | | | |
| *** Question which was not originally in the top 10 but the young peer researchers contributing throughout the study felt it was very important. | | | | |
