## Supplementary material for "Priorities for Future Research about Screen Use and Adolescent Mental Health: A Participatory Prioritization Study": YPAG Consent

1/6/2021

To whom it may concern,

We confirm Syinat Tageava has been a member of the McPin Foundation Young People's Advisory Group (YPAG) throughout this project and we have overseen her contributions, including the lived experience commentary.

Many Thanks,

Rachel K Temple

**Rachel Kimberley Temple** | Public Involvement in Research Manager | Pronouns: She/her

**Keep in touch:** Sign up to our Young People's Network here
**The McPin Foundation** | 7-14 Great Dover Street| London | SE1 4YR
