## Supplementary material for "Priorities for Future Research about Screen Use and Adolescent Mental Health: A Participatory Prioritization Study": YPAG Consent

Institute of Psychiatry, Psychology & Neuroscience

KING'S College LONDON

20/05/2021

Dear Ms. Syinat Tageava,

Thank you for your contribution to our article by co-writing the lived experience commentary.

*Please sign in the space provided below to confirm that:*

- *You are a member of the McPin Young People's Advisory Group (YPAG)*
- *You have read and understood the research study described in the paper.*
- *You have been sufficiently informed about the research methods and ethics.*
- *You gave informed consent to writing the lived experience commentary.*
- *You gave informed consent to your name appearing on the manuscript.*
- *You are 18+ years of age.*

With thanks,

Dr. Norha Vera San Juan

Signature for consent – Ms. Syinat Tageava
